# Knowledge, attitude, practice and perception regarding COVID-19 among students in Bangladesh: Survey in Rajshahi University

**DOI:** 10.1101/2020.04.21.20074757

**Authors:** Md. Abdul Wadood, ASMA Mamun, Md. Abdur Rafi, Md kamrul Islam, Suhaili Mohd, Lai Lee Lee, Md. Golam Hossain

## Abstract

**Background:** The number of infection and death by COVID-19 has been rapidly increasing since December 2019 in all over the world. Until now, there is no specific treatment or vaccine for this disease; WHO suggests only some protective measures like maintaining social distance, staying home, washing hands with soap or sanitizer, wearing mask etc. The objective of this study was to survey knowledge, attitude, practice and perception regarding COVID-19 among students in Rajshahi University, Bangladesh.

**Methods:** We collected data from 305 students of Rajshahi University for this cross-sectional study using mixed sampling from March 11 to March 19, 2020. Frequency distribution, Mann-Whitney and Kruskal-Wallis tests were used in this study.

**Results:** Out of 305 participants, 224 (73.4%) and 81 (26.6%) were male and female students respectively. The study revealed that Rajshahi university students had average knowledge on symptoms, protective way and transmission of COVID-19. Female students were more knowledgeable than male. More than one third of the students had negative attitude to avoiding public transport and going out to public places with friends and family. The practice of students practice during our data collection period and in future was not satisfactory. More than one third of students were not keen to stay at home and avoid going to crowded places. The perception towards COVID-19 was not good; they had no idea whether the outbreak would affect their daily routine, study and financial matters, study field work and restrict leisure time of meeting family and relatives.

**Conclusions:** We found that general knowledge, attitude, practice and perception of the university students regarding COVID-19 were not satisfactory. This indicated that the situation was worse among common people. In Bangladesh, the number of healthcare providers is insufficient. University students can be employed as potential workforce to create awareness among mass people on prevention of COVID-19.

## Introduction

Novel coronavirus disease 2019 (COVID-19) has appeared as one of the most severe pandemic and fatal disease in human history. It has already affected millions of people, thousands of whom are dying every day creating panic and a global deadlock in all spheres of life. The health authorities in Wuhan City of China found 27 pneumonia cases of unknown etiology on 31 December, 2019, and Wuhan’s Huanan Seafood Wholesale Market was detected as the source of the infection [1] that turned into an outbreak. On January 9, 2020, the China Government reported that the causative agent of the outbreak was ‘severe acute respiratory syndrome coronavirus 2 (SARS-CoV-2)’. The World Health Organization (WHO) named the virus ‘2019 novel coronavirus’ (2019NCV) and termed the disease ‘COVID-19’ [2]. Later, WHO declared the outbreak a Public Health Emergency of International Concern [3]. The Chinese Center for Disease Control and Prevention reported that the fatality rate of diagnosed cases was 2.3%, with an increasing risk in the subjects aged 60 and older (3.6% in subjects 60-69 years old; 8% in subjects 70-79 years old; and 14.8% in subjects aged 80 and older), and those with comorbidities (case fatality rate in healthy subjects was 0.9%) [4]. According to the Corona Virus Resource Centre of John Hopkins University and Medicine, 1,498,833 people were confirmed to contract the 2019NCV by April 9, 2020 in 184 countries and out of them, 89,435 people died [5]. The scenario is changing every day with increase in number of infection and death.

Evidences show that mammals and birds are the reservoirs of 2019NCV [6-7]. In Wuhan, the virus attacked only those people who visited the wet animal market and their contacts [8-9] indicating that person-to-person transmission is the likely route for spread of 2019NCV. The virus spreads primarily through droplets of saliva or discharge from the nose of an infected person when she/he coughs or sneezes [10]. WHO reports that the best way to prevent and slow down the transmission of 2019NCV is to be well informed about it, the disease it causes, and mode of its transmission, and suggests people to wash hands with soap or use hand sanitizers frequently, avoid touching the face, mouth, nose and eyes with unwashed or non-sanitized hands, maintaining social distance, and staying home to remain protected from the infection [10]. Individuals should also practice respiratory etiquette such as coughing and sneezing into a flexed elbow, cover mouth and nose with handkerchief or tissue paper and wearing masks to avoid spreading of the virus [10]. Touching surfaces contaminated with the virus is also a way of spread of 2019NCV that may survive on surfaces for several hours and days [11]. Till now, there is no specific vaccine or medicinal treatment for COVID-19 [10]. The disease is manifested by fever, sore throat and mild to moderate respiratory problems and most cases are cured without any special treatment [10]. It may get severe and life-threatening in people of older age and with comorbidities like cardiovascular disease, hypertension, diabetes, chronic respiratory disease, and cancer [10]. Though the outbreak of COVID-19 is a very recent occurrence, many studies have already been done on it. But, to the best of our knowledge, no study has yet been conducted on it with Bangladeshi population. In Bangladesh, COVID-19 situation is aggravating day by day. As of April 9, 2020, a total of 218 confirmed cases of COVID-19 were detected and 20 of them died [12]. This might be an underestimate, as the facilities of 2019NCV test are very limited here. Bangladesh is an over-populated country with a population density of 976/sq.km. [13], where most of its people do not maintain proper personal hygiene. The situation may turn very serious here. The government and the concerned non-government authorities have taken many measures, though too late and insufficient, to contain the outbreak. Educational institutions, offices and markets have been closed, and the whole country has been put under lock-down for more than two weeks. The government, non-government, social and professional organizations as well as the print and electronic media of the country is conducting massive publicity on mode of transmission of the virus, and sign-symptoms and measures of prevention, control and treatment of COVID-19. However, there are media observations that a considerable number of people are not following the suggestions properly. It is evidenced from previous pandemics that lack of proper knowledge about the disease is associated with negative emotion among people which can further complicate the attempts of preventing the spread of the disease [14]. Taking the perspective into our consideration, we aimed to assess knowledge, attitude, practice and perception regarding COVID-19 among the students of Rajshahi University who are assets of the nation, and thought to be more knowledgeable and conscious about contemporary happenings in home and abroad.

## Material and Methods

### Materials

This cross-sectional study was conducted from March 11 to March 19, 2020. Rajshahi University was considered as the target area, and all its students were selected as population for the study. Rajshahi University is the second largest university of Bangladesh with an approximate number of 38,300 students who come here from all over the country for higher education. Our study population comprised of non-medical students.

## Methods

### Questionnaire

We developed a questionnaire following the instructions and guidelines of WHO that had five parts: (i) general information of students, (ii) knowledge on COVID-19; it was subdivided into three portions: (a) knowledge on signs and symptoms of COVID-19, (b) knowledge on the protective ways to prevent COVID-19, (c) knowledge on COVID transmission, (iii) perception about COVID-19; it was subdivided into two portions: (a) perception towards COVID-19, and (b) perception about the impact of COVID-19, (iv) attitude towards COVID-19, and (v) practice regarding COVID-19. The draft questionnaire was sent to some experts and finally it was revised following the experts’ opinions and suggestions. We could not conduct pilot study for post-testing of this questionnaire for shortage of time.

### Sample size determination

The population size was known (34,300). In such case, the following formula would be appropriate to calculate the required sample for this study: 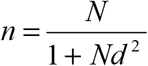, where n = required sample size, N= population size, and d= margin of error (we have considered d= 0.05) [15]. The formula provided that 396 samples would be sufficient for this study.

### Sampling and data collection

Mixed sampling was utilized in this study for selecting our required samples. In the first step, we selected three departments from 58 departments using random sampling (by lottery); the departments were: (i) Statistics, (ii) Mathematics and (iii) Physics. In the second step, one academic year was selected randomly from each selected department; these were: (i) second year from Statistics, (ii) first year from Mathematics, and (iii) first year from Physics. In the third step, cluster sampling was used for collecting information from the students of the selected years of the selected departments. Students were taken to a particular class room and briefed about the objectives of the study. Written consents of the agreed students were taken. They were abandoned from using their cell phones, laptops and internet. They filled up the questionnaire in presence of the researchers without discussion with their friends. A total number of 305 students were interviewed using this procedure. We could interview no more students as the university was closed for COVID-19 outbreak in the country. We decided to complete the study with data from 305 students.

### Ethics statement

Prior to collecting data, we took ethical clearance from the Ethical Committee, Institute of Biological Sciences (IBSc), Rajshahi University, Bangladesh to study on communicable diseases.

### Statistical analysis

All collected data were entered into SPSS (IBM, version 21) for analysis. Level of students’ knowledge on COVID-19, and percentage of their perception, attitude and practice based on the selected questions were determined by frequency distribution. The normality of the data was checked by Kolmogorov-Smirnov test. The histogram, the test and graph demonstrated that our data were not normally distributed. We used Mann-Whitney and Kruskal-Wallis (nonparametric) tests to find the significance of difference in number of correct answers between two and more than two groups respectively. Statistical significance was accepted at p < 0.05.

## Results

Of 305 samples, 73.4% were male and 26.6% were female students. Their age ranged from 17 to 28 years with mean and median ages of 20.66±1.78 and 20 years respectively. More than half of the students (51.1%) came from rural area. Near about 50% fathers and 25.6% mothers of the students were highly educated. 76.1% students came from nuclear family (Table 3). 91.8% students answered that fever was a symptom of COVID-19 followed by dry cough (81.3%), difficulty in breathing (78.4%), a general feeling of unwell (77.7%), short of breath (70.8%), headache (55.1%), running nose (54.8%), sore throat (49.8%) and diarrhea (16.4%). More than 43% reported that chest pain was also a symptom of COVID-19 (Table 2). 97.7% students believed that hand washing with soaps and water could prevent COVID-19 followed by avoidance of touching nose, mouth and eyes with unwashed hands, covering mouth and nose with tissue or handkerchief during coughing or sneezing, wearing clean surgical mask when attacked of respiratory illness (coughing, sneezing and flu), avoiding contact with people having respiratory illness symptoms such as coughing, sneezing and flu, cleaning and disinfecting objects and surface, and always eating fully cooked eggs and meat. Some students had wrong concepts also. 39.7% students believed that eating rice and vegetables could give protection from COVID-19, and another 16.1% students felt no need of maintaining distance from infected persons for protection from COVID-19 (Table 1). 97% students answered that COVID-19 could spread from person to person through coughing or sneezing, from animal to human, through objects contaminated with the virus, and eating not properly handled and cooked wild animal meat and food. More than 84% students had wrong concept that COVID-19 could transmit through touching persons with flu. 52.5% and 10.8% students believed that COVID-19 could spread through water and food, and mosquito bite respectively (Table 1).

**Table 1:**
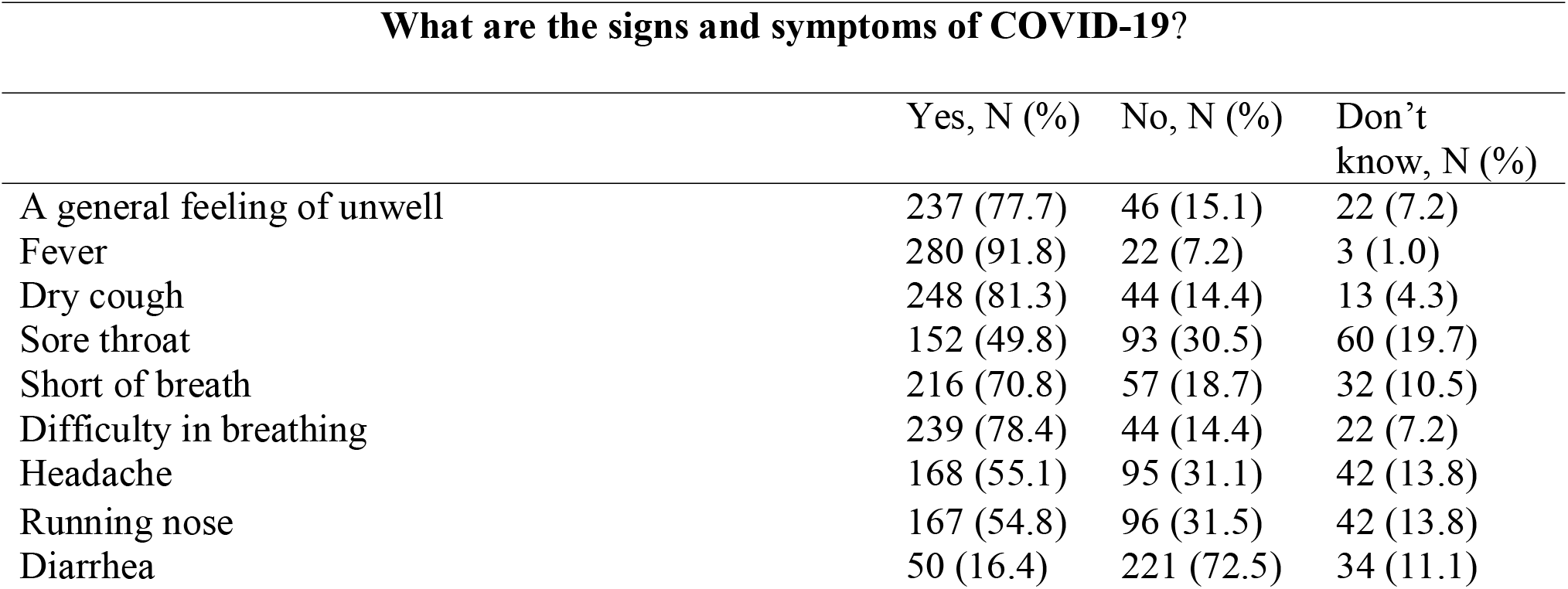
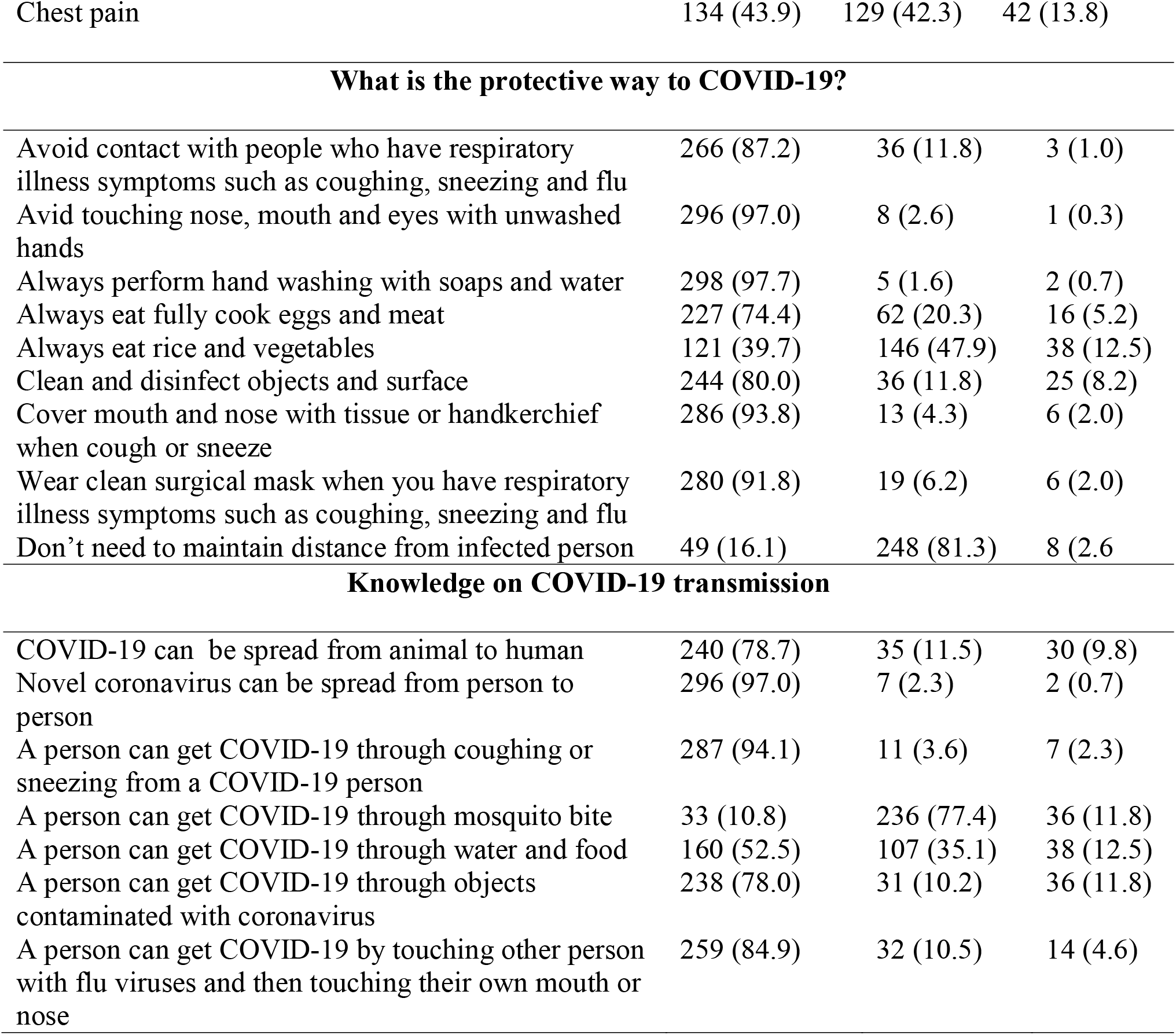
General knowledge on COVID-19

| What are the signs and symptoms of COVID-19? |  |  |  |
| --- | --- | --- | --- |
|  | Yes, N (%) | No, N (%) | Don't know, N (%) |
| A general feeling of unwell | 237 (77.7) | 46 (15.1) | 22 (7.2) |
| Fever | 280 (91.8) | 22 (7.2) | 3 (1.0) |
| Dry cough | 248 (81.3) | 44 (14.4) | 13 (4.3) |
| Sore throat | 152 (49.8) | 93 (30.5) | 60 (19.7) |
| Short of breath | 216 (70.8) | 57 (18.7) | 32 (10.5) |
| Difficulty in breathing | 239 (78.4) | 44 (14.4) | 22 (7.2) |
| Headache | 168 (55.1) | 95 (31.1) | 42 (13.8) |
| Running nose | 167 (54.8) | 96 (31.5) | 42 (13.8) |
| Diarrhea | 50 (16.4) | 221 (72.5) | 34 (11.1) |
| Chest pain | 134 (43.9) | 129 (42.3) | 42 (13.8) |
| **What is the protective way to COVID-19?** |  |  |  |
| Avoid contact with people who have respiratory illness symptoms such as coughing, sneezing and flu | 266 (87.2) | 36 (11.8) | 3 (1.0) |
| Avoid touching nose, mouth and eyes with unwashed hands | 296 (97.0) | 8 (2.6) | 1 (0.3) |
| Always perform hand washing with soaps and water | 298 (97.7) | 5 (1.6) | 2 (0.7) |
| Always eat fully cook eggs and meat | 227 (74.4) | 62 (20.3) | 16 (5.2) |
| Always eat rice and vegetables | 121 (39.7) | 146 (47.9) | 38 (12.5) |
| Clean and disinfect objects and surface | 244 (80.0) | 36 (11.8) | 25 (8.2) |
| Cover mouth and nose with tissue or handkerchief when cough or sneeze | 286 (93.8) | 13 (4.3) | 6 (2.0) |
| Wear clean surgical mask when you have respiratory illness symptoms such as coughing, sneezing and flu | 280 (91.8) | 19 (6.2) | 6 (2.0) |
| Don't need to maintain distance from infected person | 49 (16.1) | 248 (81.3) | 8 (2.6) |
| **Knowledge on COVID-19 transmission** |  |  |  |
| COVID-19 can be spread from animal to human | 240 (78.7) | 35 (11.5) | 30 (9.8) |
| Novel coronavirus can be spread from person to person | 296 (97.0) | 7 (2.3) | 2 (0.7) |
| A person can get COVID-19 through coughing or sneezing from a COVID-19 person | 287 (94.1) | 11 (3.6) | 7 (2.3) |
| A person can get COVID-19 through mosquito bite | 33 (10.8) | 236 (77.4) | 36 (11.8) |
| A person can get COVID-19 through water and food | 160 (52.5) | 107 (35.1) | 38 (12.5) |
| A person can get COVID-19 through objects contaminated with coronavirus | 238 (78.0) | 31 (10.2) | 36 (11.8) |
| A person can get COVID-19 by touching other person with flu viruses and then touching their own mouth or nose | 259 (84.9) | 32 (10.5) | 14 (4.6) |

**Table 2:** Level of knowledge on COVID-19 among university students

| Question | Number of correct answer | Level of knowledge, N (%) | Question | Number of correct answer | Level of knowledge, N (%) |
| --- | --- | --- | --- | --- | --- |
| What are the signs and symptoms of Covid-19? | 0 | 3 (1.0) | What is the protective way to avoid COVID infections? | 2 | 2 (0.3) |
|  | 1 | 5 (1.6) |  | 3 | 5 (1.6) |
|  | 2 | 7 (2.3) |  | 4 | 14 (4.6) |
|  | 3 | 13 (4.3) |  | 5 | 37 (12.1) |
|  | 4 | 36 (11.8) |  | 6 | 97 (31.8) |
|  | 5 | 53 (17.4) |  | 7 | 151 (49.5) |
|  | 6 | 82 (26.9) | Knowledge on COVID-19 transmission | 1 | 1 (0.3) |
|  | 7 | 62 (20.3) |  | 2 | 6 (2.0) |
|  | 8 | 32 (10.5) |  | 3 | 31 (10.2) |
|  | 9 | 12 (3.9) |  | 4 | 56 (18.4) |
|  |  |  |  | 5 | 104 (34.1) |
|  |  |  |  | 6 | 107 (35.1) |

**Table 3:** Average knowledge on symptoms, protective way and transmission of COVID-19 by different characteristics of participants

| Characteristics | Sings and symptoms, Mean±SD, $5.76 \pm 1.76$ | Protective way to avoid COVID-19, Mean±SD, $6.22 \pm 0.98$ | Knowledge on transmission, Mean±SD, $4.89 \pm 1.08$ |
| --- | --- | --- | --- |
| **Gender** | M-W test value=8173<sup>n</sup> | M-W test value=7587<sup>*</sup> | M-W test value=7534<sup>*</sup> |
| Male, 224(73.4) | $5.68 \pm 1.79$ | $6.11 \pm 1.05$ | $4.80 \pm 1.11$ |
| Female, 81(26.6) | $5.99 \pm 1.66$ | $6.48 \pm 0.67$ | $5.14 \pm 0.96$ |
| **Original residence** | M-W test value=11070<sup>n</sup> | M-W test value=11128<sup>n</sup> | M-W test value=11570<sup>n</sup> |
| Urban, 149(48.9) | 5.69±1.75 | 6.25±0.98 | 4.91±1.01 |
| Rural, 156(51.1) | 5.83±1.77 | 6.19±0.98 | 4.87±1.15 |
| **Type of family** | M-W test<br>value=79430<sup>n</sup> | M-W test<br>value=8283<sup>n</sup> | M-W test<br>value=0.948<sup>n</sup> |
| Nuclear, 232(76.1) | 5.79±1.77 | 6.20±1.00 | 4.89±1.07 |
| Joint,<br>73(23.9) | 5.67±1.72 | 6.27±0.90 | 4.89±1.11 |
| **Fathers' education** | K-W test,<br>Chi-square<br>value=1.074<sup>n</sup> | K-W test,<br>Chi-square<br>value=1.712<sup>n</sup> | K-W test,<br>Chi-square<br>value=5.372<sup>n</sup> |
| Uneducated, 25(8.2) | 5.28±2.23 | 6.12±0.93 | 5.28±0.79 |
| Primary, 48(15.7) | 5.79±1.69 | 6.08±1.15 | 4.71±1.11 |
| Secondary, 86(28.2) | 5.92±1.69 | 6.28±0.99 | 4.80±1.09 |
| Higher, 146(47.9) | 5.74±1.73 | 6.25±0.92 | 4.94±1.09 |
| **Mothers' education** | K-W test,<br>Chi-square<br>value=3.704<sup>n</sup> | K-W test,<br>Chi-square<br>value=0.832<sup>n</sup> | K-W test,<br>Chi-square<br>value=3.228<sup>n</sup> |
| Uneducated, 36(11.8) | 5.22±2.14 | 6.17±0.91 | 4.83±1.06 |
| Primary, 65(21.3) | 5.94±1.60 | 6.18±0.97 | 4.69±1.16 |
| Secondary, 126(41.3) | 5.87±1.76 | 6.22±1.00 | 4.95±1.09 |
| Higher, 78(25.6) | 5.67±1.67 | 6.27±0.99 | 4.99±1.00 |

**N.B.:** SD: Standard deviation; M-W: Mann-Whitney; K-W: Kruskal-Wallis; \*: 5% level of significance; n: non-significance.

Table 2 shows the level of knowledge on COVID-19 among university students. Only 3.9% students gave correct answers of all questions regarding signs and symptoms of COVID-19, while more than 46% students answered 6-7 questions correctly. About half of the students (49.5%) knew the ways of prevention of COVID-19. 35.1% students had knowledge about the possible ways of COVID-19 transmission.

The average number of 5.76±1.76 questions on signs and symptoms, 6.22±0.98 questions on protective way to avoid COVID-19 and 4.89±1.08 questions on transmission of COVID-19 were correctly answered by the participants. Mann-Whitney test demonstrated that female students gave significantly more correct answers to the questions regarding protective way to avoid and transmission of COVID-19 than male students (p<0.05). No significant difference was observed between urban and rural, nuclear and joint family, and among parent’s education levels regarding knowledge on COVID-19 (Table 3).

82.0% students were afraid to contact people with flu symptoms followed by eating wildlife animal’s meat, going abroad with friends and family, eating raw food, going to crowed places, eating outside food from hawker centers, contacting friends and relatives just coming back from overseas, going out to public places with friends and family, and taking public transport. On the other hand, 41.6% and 30.5% did not like to avoid taking public transport and going out to public places with friends and family respectively (Table 4).

**Table 4:**
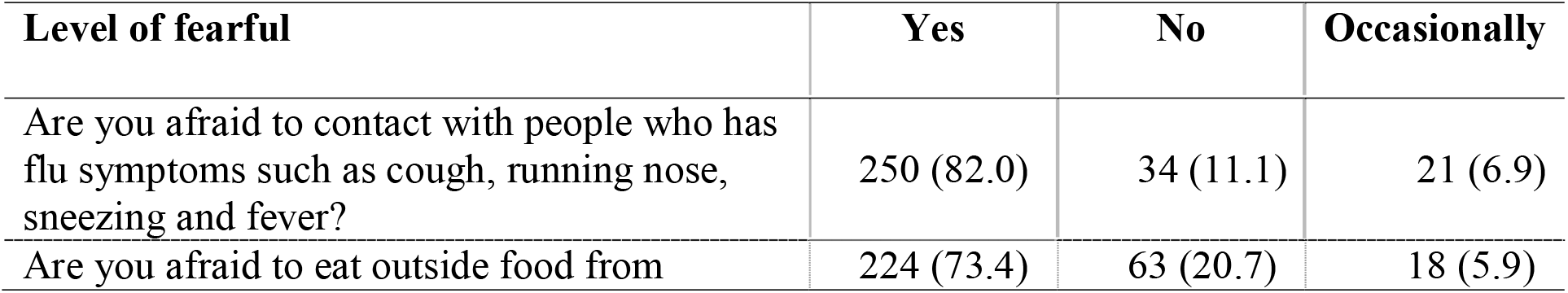
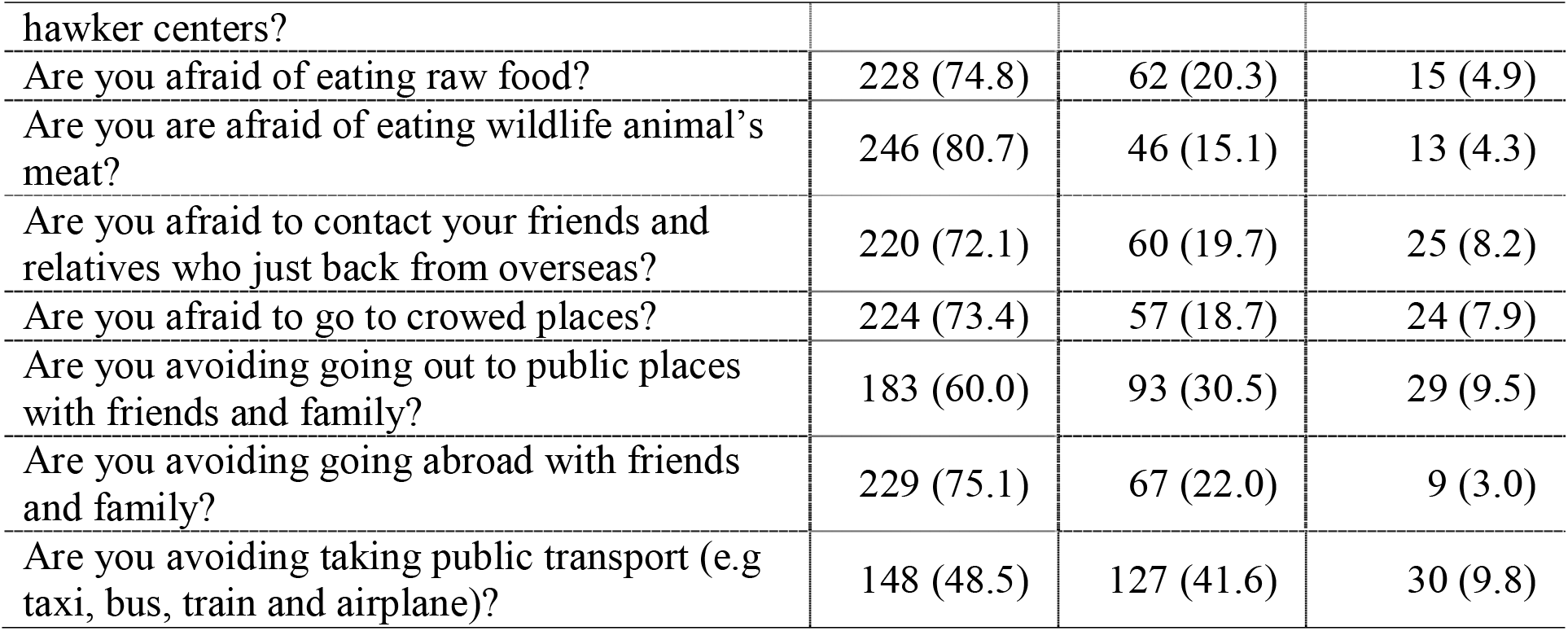
Attitude toward COVID-19 among university students

It was observed that 89.5%, 82.3%, 66.6%, 59.7% and 53.8% students currently had or will have the practice of washing hands frequently, performing healthy lifestyle, taking medicines if necessary, staying home and avoiding crowded places and wearing surgical face masks while going out in public places respectively to avoid COVID-19 attack. More than 39% students had negative practice of not wearing surgical face masks while going out in public places, and 32.1% and 27.2 % students will not like to practice staying at home and avoid going to crowded places and taking medicine if they feel unwell respectively (Table 5).

**Table 5:** Practice toward COVID-19 among university students

| **Practice to avoid COVID-19** | **Yes** | **No** | **Occasionally** |
| --- | --- | --- | --- |
| Do you wash your hands frequently and thoroughly? | 273 (89.5) | 18 (5.9) | 14 (4.6) |
| Are you are wearing surgical face mask when out in public? | 164 (53.8) | 121 (39.7) | 20 (6.6) |
| Will you stay at home and avoid go to crowded places? | 182 (59.7) | 98 (32.1) | 25 (8.2) |
| Will you take medicine (conventional or traditional medicine) if you feel your body unwell? | 203 (66.6) | 83 (27.2) | 19 (6.2) |
| Will you perform healthy lifestyle (eat nutritious food, exercise regularly, and get enough of rest or sleep) to maintain your body health and to avoid infection? | 251 (82.3) | 32 (10.5) | 22 (7.2) |

Only 21% students showed positive attitude answering all 9 questions correctly, while 36.4%, 29.2% and 12.5% students showed positive attitude with correctly answering 7-8, 4-6 and 1-3 questions respectively. Proper practices (answering all questions correctly) were found among 24% students while 54.1% and 20.4% students provided 3-4 and 1-2 correct answers showing their poor practice (Table 6).

**Table 6:** Level of positive attitude and good practice on COVID-19 among university students

| Attitude | Number of positive answer | Level of positive attitude, N (%) | Practice | Number of positive answer | Level of good practice, N (%) |
| --- | --- | --- | --- | --- | --- |
| Attitude toward COVID-19 | 0 | 3 (1.0) | Practice toward COVID-19 | 0 | 3 (1.0) |
|  | 1 | 5 (1.6) |  | 1 | 13 (4.3) |
|  | 2 | 24 (7.9) |  | 2 | 49 (16.1) |
|  | 3 | 9 (3.0) |  | 3 | 73 (23.9) |
|  | 4 | 22 (7.2) |  | 4 | 92 (30.2) |
|  | 5 | 5 (10.5) |  | 5 | 75 (24.6) |
|  | 6 | 35 (11.5) |  |  |  |
|  | 7 | 50 (16.4) |  |  |  |
|  | 8 | 61 (20.0) |  |  |  |
|  | 9 | 64 (21.0) |  |  |  |

The average level of positive attitude and good practice among students were 6.40±2.32 and 3.52±1.20 respectively. Mann-Whitney test demonstrates that female students (3.86±1.16) were significantly more practicing to avoid COVID-19 than male students (3.39±1.19) (p<0.01), and students coming from rural environment (6.70±2.12) showed more positive attitude than their counterparts (6.09±2.49) (p<0.05). Students living in nuclear family (3.62±1.16) had more practice to avoid COVID-19 than students coming from joint family (3.21±1.28) (p<0.05). We did not find the significance of difference in attitude and practice between other groups (Table 7).

**Table 7:** Positive attitude and good practice of COVID-19 by different characteristics of participants

| **Characteristics** | **Attitude , Mean±SD, 6.40±2.32** | **Practice, Mean±SD, 3.52±1.20** |
| --- | --- | --- |
| **Gender** | M-W test value=8015<sup>n</sup> | M-W test value=6964<sup>**</sup> |
| Male, 224(73.4) | 6.33 ±2.21 | 3.39±1.19 |
| Female, 81(26.6) | 6.58±2.62 | 3.86±1.16 |
| **Original residence** | M-W test value=10110<sup>*</sup> | M-W test value=10868<sup>n</sup> |
| Urban, 149(48.9) | 6.09±2.49 | 3.46±1.15 |
| Rural, 156(51.1) | 6.70±2.12 | 3.57±1.25 |
| **Type of family** | M-W test value=8376<sup>n</sup> | M-W test value=6935<sup>*</sup> |
| Nuclear, 232(76.1) | 6.42±2.29 | 3.62±1.16 |
| Joint, 73(23.9) | 6.33±2.43 | 3.21±1.28 |
| **Fathers' education** | K-W test, $\chi^2$-value=6.607<sup>n</sup> | K-W test, $\chi^2$-value =5.009<sup>n</sup> |
| Uneducated, 25(8.2) | 7.00±1.78 | 3.24±1.23 |
| Primary, 48(15.7) | 6.94±2.39 | 3.83±1.16 |
| Secondary, 86(28.2) | 6.24±2.44 | 3.49±1.23 |
| Higher, 146(47.9) | 6.21±2.28 | 3.48±1.19 |
| **Mothers' education** | K-W test, $\chi^2$-value=3.507<sup>n</sup> | K-W test, $\chi^2$-value=2.758<sup>n</sup> |
| Uneducated, 36(11.8) | 6.78±2.09 | 3.22±1.38 |
| Primary, 65(21.3) | 6.65±2.43 | 3.68±1.17 |
| Secondary, 126(41.3) | 6.38±2.19 | 3.52±1.21 |
| Higher, 78(25.6) | 6.05±2.52 | 3.51±1.13 |

**N.B.:** SD: Standard deviation; M-W: Mann-Whitney; K-W: Kruskal-Wallis; $\chi^2$ : Chi-square; \*: 5% level of significance; \*\*: 1% level of significance; n: non-significance.

54.1% students felt they were highly exposed to COVID-19, while near about half of the students thought they were not or not at all exposed. More than 36%, 30%, 27% and 6% students were extremely, very, moderately and not at all worried about getting COVID-19 respectively. 36.4% and 27.5% students were very and moderately worried about the consequences of getting COVID-19 respectively. About one-third (32.5%), 37.7% and 22.0% students were very fearful, fearful and slightly fearful respectively of COVID-19. 23%, 27.5%, 26.6% and 23% students felt that COVID-19 outbreak affected their daily routine with great extent, moderately, very little and not at all respectively. Also, 23%, 17%, 22.3% and 37.7% students thought COVID-19 outbreak affected their study with great extent, moderately, very little and not at all respectively. More than 21%, 16%, 19% and 43% students believed that COVID-19 affected financial matters with great extent, moderately, very little and not at all respectively. More than 29% students did not think that COVID-19 outbreak affected their family’s daily routine, while 18%, 30.2% and 22.6% students believed that it affected their family’s daily routine with great extent, moderately and very little respectively. About 60% students answered that COVID-19 affected their travel abroad with great extent, while other students thought it had affected their travel abroad moderately (17.4%), little (11.8%) or not at all (11.5%). More than 23% students did not believe that COVID-19 affected their study field works, while more than 33% and 22% students thought it affected their study field works with great extent and moderately respectively. Students believed with great extent (31.1%), moderate (23%), very little (26.9%) and not at all (19%) that COVID-19 restricted their leisure time of meeting friends. They also believed that COVID-19 outbreak restricted their leisure time of meeting family and relatives with great extent (28.9%), moderately (23%), very little (18%) and not at all (30.2%) (Table 8).

**Table 8:** Perception of students toward COVID-19 and about its impact

|  |  |  |  |  |
| --- | --- | --- | --- | --- |
| Are you highly exposed to COVID-19? | Yes, N (%)<br>165 (54.1) | No, N (%)<br>79 (25.9) | Not at all, N (%)<br>61 (20.0) |  |
|  | Extremely<br>N (%) | Very<br>N (%) | Moderately<br>N (%) | Not at all<br>N (%) |
| How worried are you about getting COVID-19? | 110 (36.1) | 92 (30.2) | 84 (27.5) | 19 (6.2) |
| How worried are you about the consequences of getting COVID-19? | 96 (31.5) | 111 (36.4) | 84 (27.5) | 14 (4.6) |
| How is your fear level toward COVID-19? | Very fearful<br>N (%)<br>99 (32.5) | Fearful<br>N (%)<br>115 (37.7) | Slightly fearful<br>N (%)<br>67 (22.0) | Not at all fearful,<br>N (%)<br>24 (7.9) |

Perception about the impact of COVID-19
|  | Great extent | Moderately | Very little | Not at all |
| --- | --- | --- | --- | --- |
| The COVID-19 outbreak has affected my daily routine | 70(23.0) | 84(27.5) | 81(26.6) | 70(23.0) |
| The COVID-19 outbreak has affected my study | 70(23.0) | 52(17.0) | 68(22.3) | 115(37.7) |
| The COVID-19 outbreak has affected my financial | 65(21.3) | 49(16.1) | 60(19.7) | 131(43.0) |
| The COVID-19 outbreak has affected my family's daily routine | 55(18.0) | 92(30.2) | 69(22.6) | 89(29.2) |
| The COVID-19 outbreak has affected my travel abroad | 181(59.3) | 53(17.4) | 36(11.8) | 35(11.5) |
| The COVID-19 outbreak has affected my study field work | 102(33.4) | 69(22.6) | 63(20.7) | 71(23.3) |
| The COVID-19 outbreak has restrict my leisure time of meeting friends | 95(31.1) | 70(23.0) | 82(26.9) | 58(19.0) |
| The COVID-19 outbreak has restrict my leisure time of meeting family and relatives | 88(28.9) | 70(23.0) | 55(18.0) | 92(30.2) |

## Discussion

The outbreak of COVID-19 in China attracted attention of the whole world including Bangladesh since its early stage. Hundreds of students, businessmen and tourists started to return home from China creating a hue and cry in the country. Soon Bangladeshi people from many other countries started to rush to the homeland. Mismanagement of health check-up, COVID-19 test and 14 days quarantine of these home-coming people created a huge debate and became a hot cake for print, electronic and social media. Being comparatively more knowledgeable part of the population, the university students are supposed to have higher level of knowledge, attitude, practice and perception regarding the disease. For that purpose, data were collected from 305 students of Rajshahi University. It is reported that adequate knowledge, and positive attitude and perception are related to proper practice [16]. Four types of issues related to COVID-19 were discussed in this study.

### Knowledge on COVID-19

### Signs and symptoms

This study revealed that only 3.9% and 10.5% students knew correct answers of all nine and eight questions respectively. This indicates that the students did not have good knowledge about the signs and symptoms of COVID-19. The knowledge level of our students was far below than that of Chinese people [2] and Indian healthcare professionals, students and non-medical healthcare staffs [17].

#### Protective way to avoid COVID-19

About 40% students thought eating rice and vegetables could prevent COVID-19. Half of the students answered all seven questions correctly. We may consider that the students had good knowledge on the protective ways of avoiding COVID-19 though the level of knowledge was lower than that among Indian healthcare professionals, students and non-medical healthcare staffs [17].

#### Knowledge of COVID-19 transmission

More than half of the students deemed that COVID-19 could transmit via water and food, and 10.8% students believed that mosquito was a vector of COVID-19. Only 35.1% students answered all the 6 questions correctly. It reveals that the students had fair or moderate knowledge on COVID-19 transmission. However, it was lower than the knowledge level of healthcare professionals and students but higher than that among non-medical healthcare staffs of India [17]. We found that female students were more knowledgeable than males regarding transmission of COVID-19. Usually females take everything more seriously than males, and they have more curiosity to know anything than males. This might be a reason of difference of the knowledge between male and female students. Same result was observed in Chinese and Indian studies [16, 17].

### Attitude

WHO suggests people to avoid public gatherings, maintain social distancing and stay home for prevention of COVID-19. This study demonstrates that more than 40% had negative attitude towards the measures. The facts reveal that the students of Rajshahi University did not have good attitude towards COVID-19. Attitude towards COVID-19 among the Chinese people and Indian healthcare professionals, students and non-medical healthcare staffs was very positive [2, 17]. The students coming from rural area showed more positive attitude than urban students. It could not be compared as no similar study is available.

### Practice

About 40% students were not using surgical face masks in public places. Scarcity and higher price of masks might be a reason behind it. More than one-third of students were not interested in staying home and avoiding crowds. In China, almost all people used face masks and avoided public places to prevent COVID-19 transmission [2]. Level of practice of preventive guides was also high among Indians [17]. Usually, practice of personal hygiene is comparatively poor in Bangladesh, such as, only 40% people wash hands with soap and water and only 35% schools have facilities of washing hands with soap and water [17]. This might contribute to poor practice regarding prevention of COVID-19 among our respondents. The female students coming from nuclear family were more practicing to avoid COVID-19 than males and joint family’s students. A Chinese study supports the finding [16].

### Perception

#### Perception towards COVID-19

A considerable number of students thought that they were not exposed to COVID-19. Only a smaller part of them were moderately worried of its infection and consequences, and slightly fearful of it. On the basis of answers of the four questions of this section, we can conclude that the student’s perception towards COVID-19 was not good. No previous study on this issue is available till now to compare our finding. Religious and cultural faiths and views might play role in this regard.

#### Perception about the impact of COVID-19

This study shows that only a smaller portion of students felt COVID-19 outbreak affected their daily routine, study, financial matters, family’s daily routine, study field works, and leisure time. The respondents’ perception regarding impact of COVID-19 was poor. To the best of our knowledge, there is no relevant study either in home or abroad regarding the perception about the impact or consequences of COVID-19. We think, during our data collection period, students had no idea how COVID-19 could affect their life.

#### Limitation of this study

Perhaps, this is the first attempt of survey on knowledge, attitude, practice and perception towards COVID-19 among university students in Bangladesh. This study searched out some alarming facts. However, we had many limitations too. The sample size was small. This cross-sectional study could not look into any change in students’ knowledge, attitude, practice and perception on COVID-19 in course of time. By this time, some new information was got, and new misconceptions and misinformation were publicized, but we could not consider these issues. There are many universities in Bangladesh, but we considered only Rajshahi University students as our sample due to limitation of time and budget. The limitations reveal that more in-depth studies are needed.

## Conclusions

In this study, we examined the students’ knowledge on signs and symptoms, preventive measure and mode of transmission of COVID-19. We found that the general knowledge of the students on our selected issues was not satisfactory. A remarkable number of students had negative attitude towards COVID-19. During the data collection time, students’ practices of avoiding COVID-19 were found unsatisfactory. Also, students’ perception about COVID-19 was not good. We may suggest the health authorities of Bangladesh to take necessary steps for improving university students’ knowledge, attitude, practice and perception towards COVID-19. In our country where the number of doctors and other healthcare providers are not sufficient, knowledgeable university students can be employed to create awareness on COVID-19 among mass people.

## Data Availability

Data is available. If requested, we can provide it.

## Acknowledgements

The authors gratefully acknowledge the authority of the selected departments, University of Rajshahi, Bangladesh for providing information about the students. The authors would also like to express their sincere gratitude to all the participants for proving their information.

## References

1. Wuhan Municipal Health and Health Commission’s briefing on the current pneumonia epidemic situation in our city 2019. Available online: http://wjw.wuhan.gov.cn/front/web/showDetail/2019123108989 (accessed on 27 March, 2020).

2. Naming the coronavirus disease (World Health Organization). Available online: https://www.who.int/emergencies/diseases/novel-coronavirus-2019/technical-guidance/naming-the-coronavirusdisease-(covid-2019)-and-the-virus-that-causes-it (accessed on 27 March, 2020).

3. Sohrabia C, Alsafib Z, O’Neilla N, Khanb M, Kerwanc A, Al-Jabirc A, Iosifidisa C, Aghad R. World Health Organization declares global emergency: A review of the 2019 novel coronavirus (COVID-19). International Journal of Surgery, 2020; 76:71–76

4. The Novel Coronavirus Pneumonia Emergency Response Epidemiology Team, Weekly Surveillance (Chinese Centers for Disease Control and Prevention). Available online: http://weekly.chinacdc.cn/en/article/id/e53946e2-c6c4-41e9-9a9b-fea8db1a8f51 (accessed on 27 March, 2020).

5. Corona Virus Resource Centre, John Hopkins University and Medicine. Available online: https://coronavirus.jhu.edu/map.html (accessed on April 9, 2020).

6. M. Bassetti, A. Vena, D. Roberto Giacobbe, The Novel Chinese Coronavirus (2019- nCoV) Infections: challenges for fighting the storm, Eur. J. Clin. Invest. 2020; e13209. https://doi.org/10.1111/eci.13209.

7. W. Ji, W. Wang, X. Zhao, J. Zai, X. Li, Homologous recombination within the spike glycoprotein of the newly identified coronavirus may boost cross-species transmission from snake to human, J. Med. Virol. 2020; 92 (4):433–440. https://doi.org/10.1002/jmv.25682.

8. W.G. Carlos, C.S. Dela Cruz, B. Cao, S. Pasnick, S. Jamil, Novel wuhan (2019-nCoV) coronavirus, Am. J. Respir. Crit. Care Med. 2020; 201 (4):7–8. https://doi.org/10.1164/rccm.2014P7.

9. P. Wu, X. Hao, E.H.Y. Lau, J.Y. Wong, K.S.M. Leung, J.T. Wu, et al., Real-time tentative assessment of the epidemiological characteristics of novel coronavirus infections in Wuhan, China, as at 22 January 2020. Euro Surveill. 2020; 25.

10. World Health Organization, Corona Virus. https://www.who.int/health-topics/coronavirus#tab=tab_1 (accessed on 27 March, 2020).

11. UNICEF. Coronavirus disease (COVID-19): What parents should know. Available online: https://www.unicef.org/bangladesh/en/coronavirus-disease-covid-19-what-parents-should-know.

12. World Health Organization, Corona Virus Disease (COVID-19) Outbreak. Available online: https://www.who.int/southeastasia/outbreaks-and-emergencies/novel-coronavirus-2019 (accessed on April 9, 2020).

13. Complimentary population monograph of Bangladesh. Bangladesh Bureau of Statistics (BBS), Dhaka, November 2015. Available online: http://203.112.218.65:8008/WebTestApplication/userfiles/Image/PopMonographs/Volume-7_PDV.pdf (Accessed on April 6, 2020).

14. Person B, Sy F, Holton K, Govert B, Liang A, Garza B, et al. Fear and Stigma: The Epidemic within the SARS Outbreak. Vol. 10, Emerging Infectious Diseases. Centers for Disease Control and Prevention (CDC); 2004; 358–63.

15. Rana M, Sayem A, Karim R, Islam N, Islam R, Zaman TK, Hossain G. Assessment of knowledge regarding tuberculosis among non-medical university students in Bangladesh: a cross-sectional study. BMC Public Health. 2015; 15:716. doi: 10.1186/s12889-015-2071-0.

16. Zhong B-L, Luo W, Li H-M, Zhang Q-Q, Liu X-G, Li W-T, et al. Knowledge, attitudes, and practices towards COVID-19 among Chinese residents during the rapid rise period of the COVID-19 outbreak: a quick online cross-sectional survey. Int. J. Biol. Sci. 2020; 16(10): 1745–1752. doi: 10.7150/ijbs.45221.

17. Modi PD, Nair G, Uppe A, Modi J, Tuppekar B, Gharpure AS and Langade D. COVID-19 Awareness among Healthcare Students and Professionals in Mumbai Metropolitan Region: A Questionnaire Based Survey. Cureus, 2020; 12(4): e7514.

